# Non-binary gender identity associates with decreased functioning during an exceptional societal stress. A university community study during COVID-19 pandemic in Finland

**DOI:** 10.64898/2026.01.10.25343131

**Authors:** Raimo K. R. Salokangas, Tiina From, Jarmo Hietala

## Abstract

**Background and Aim:** Individuals with non-binary gender commonly face minority stress such as invisibility, identity invalidation or even hostility within various social contexts. The COVID-19 pandemic represented an exceptional form of societal stress that presented mental health challenges in the population and probably even more so for vulnerable groups including non-binary gender individuals. We investigated whether there are gender differences in the impact of the COVID-19 pandemic on functional ability.

**Method:** University students and personnel (n=1998) responded to an online survey in May 2021, when the measures for preventing COVID-19 infections had sustained about a year and a half. Based on the gender option responses, groups of non-binary and binary (male or female) gender identity were formed. Current functioning (FUNCT), and subjective assessment of the effect of COVID-19 on functioning (COFUNCT) were recorded. Psychosocial and mental health characteristics were included in the statistical models.

**Results:** The non-binary group represented 3,6 % of all study participants. The gender option “Male” was selected by 23.8% and the gender option “Female” by 72.7% of respondents. Compared to the binary group, those in the non-binary group exhibited poorer socioeconomic living situation and less favourable previous psychosocial development. Non-binary participants reported lower FUNCT and more negative COFUNCT than binary participants. In non-binary participants, a poor work situation was directly associated with poor FUNCT, while multiple adverse childhood experiences and loneliness were indirectly associated with lower FUNCT via depressive symptoms. Conversely, high family support and previous mental health care were directly associated with more negative COFUNCT, and loneliness was indirectly associated with low COFUNCT via depression. In binary participants, family support, good economy, resilience and active physical exercise associated with good FUNCT, while age, family support, good economy, resilience, active physical exercise and adverse childhood experiences associated with good COFUNCT.

**Conclusions:** Individuals with non-binary gender are more vulnerable for functional deficits in a period of serious societal stress such as COVID-19 pandemic. The related psychosocial and mental health factors should be taken into account when planning tailored interventions for vulnerable groups during periods of exceptional societal circumstances.

**Graphical abstract:** 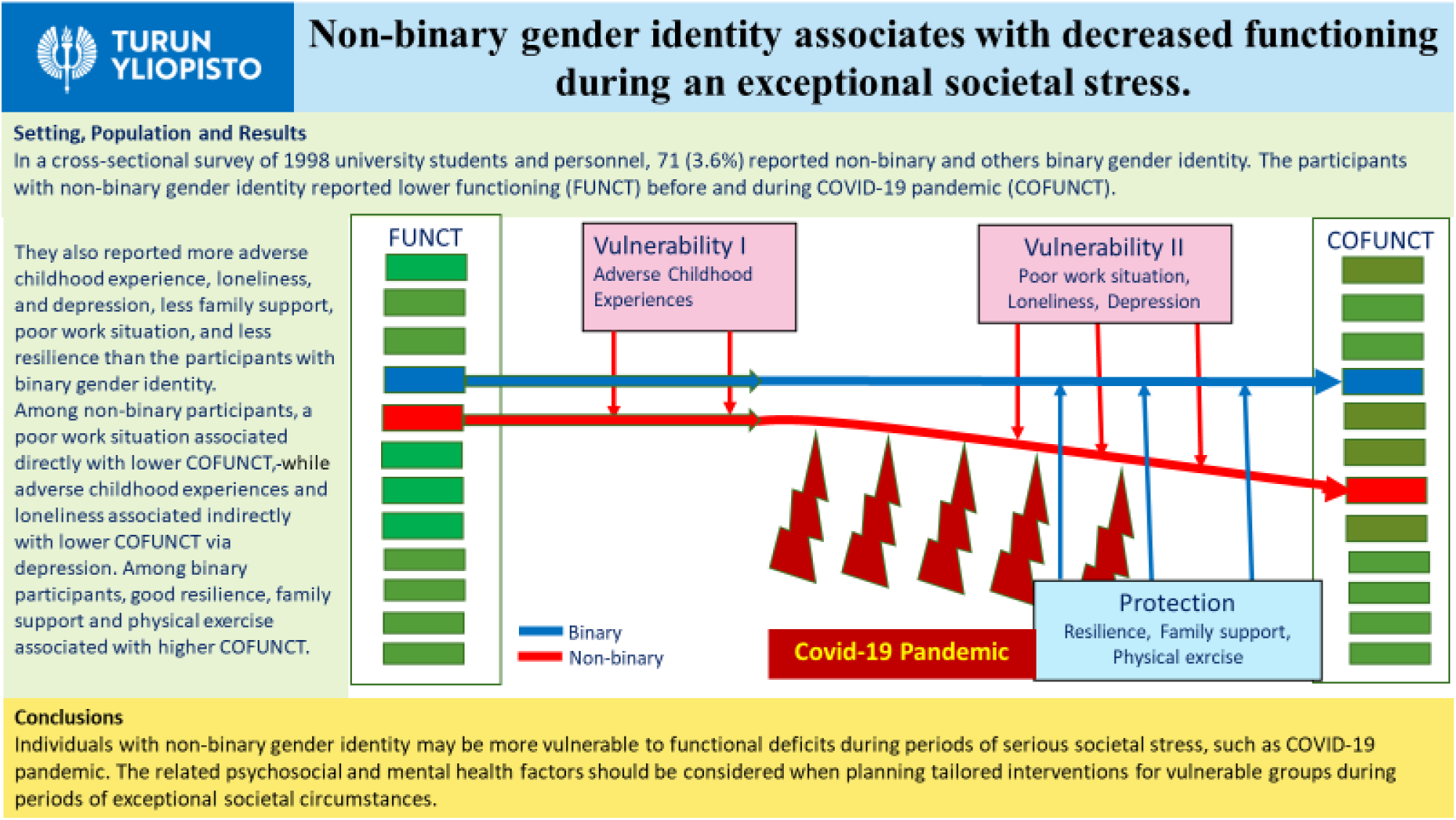

## 1. Introduction

### 1.1. Non-binary gender

Non-binary or genderqueer identity is a commonly used umbrella term to describe identities that exist outside of the binary concept of gender [1,2]. People who identify as non-binary may feel that their gender is a mix of both, neither, or something entirely different, including but not to limited to genderqueer, genderfluid, agender, bigender, and transgender [3]. It’s important to note that non-binary gender identity is distinct from sexual orientation and can vary widely from person to person.

In population surveys, non-binary identity ranges from 0.3% to 0.5% among adults, and from 1.2% to 2.7% among children and adolescents. Using a broader concept of gender diversity, the corresponding proportions increased to 0.5-4.5% among adults, and 2.5-8.4% among children and adolescents [4]. Although not being disorder, non-binary individuals are still at risk of experiencing minority stress or marginalization [3]. Non-binary individuals often find themselves on the fringes of society, where they encounter significant challenges such as stigma, discrimination, exclusion, and violence, leading to poorer health outcomes [5]. Consequently, they are at a higher risk of experiencing psychological distress, e.g., depression, anxiety, and suicidality, than their cisgender heterosexual peers [6]. In the elderly population, non-binary individuals are associated with poorer physical and mental health, and they are more likely to violence and loneliness [7]. Among clinically-referred adolescents and adults, identifying more strongly with a non-binary identity was associated with poorer psychological functioning [8]. All these vulnerability factors may limit non-binary individuals’ ability to work and function in normal societal circumstances; even more when a society has faced an exceptional stress situation like COVID-19 pandemic.

### 1.2. The COVID-19 pandemic as a societal stress

The combination of the COVID-19 pandemic and measures for preventing COVID-19 infections led to high levels of social and personal stress [9,10], thereby resulting in social isolation, subjective feelings of loneliness, and a high likelihood of psychological distress symptoms, such as depression and anxiety [11,12]. In the beginning of the COVID-19 pandemic, high levels of resilience, active physical exercise, and strong social support from family and friends were associated with fewer symptoms of distress [13,14].

Population surveys, carried out during COVID-19 pandemic, found that e.g., that young age, non-binary gender, and low levels of resilience have been associated with symptoms of psychological distress [15,16,17]. Societal stress related to the COVID-19 pandemic, most strongly affected socially or economically marginalised groups, including low-income individuals, women, transgender people, children, those with a history of childhood adversity, and socially isolated and lonely people [18]. Consequently, it can be assumed that individuals with non-binary gender, experiencing marginalisation stress during the COVID-19 pandemic, faced compounded stress.

The COVID-19 pandemic had a strong effect on university communities via lockdowns, remote education, and bans on gatherings, resulting in decreased wellbeing, loneliness, anxiety, depression and lower levels of academic success [19–22]. Hence, a university community offers a good environment for studying effects of societal stress. Moreover, within the university community, individuals with non-binary gender often face social discrimination, poly-victimization, and psychological and/or physical abuse [23].

The present study is a secondary data analysis based on our previous study on mental health among university population during the COVID-19 pandemic [24]. In this study, we aimed to investigate how individuals’ gender identity associates with everyday functioning during an exceptional societal stress (COVID-19 pandemic). We hypothesized that the individuals with non-binary gender report lower functioning (FUNCT) and more negative effect of COVID-19 on functioning (COFUNCT) than the individuals with binary gender, and binary males and binary females separately. We first compare sociodemographic background and other descriptive (confounding) factors of individuals with the gender identity. Secondly, we compare FUNCT and COFUNCT between participants with different gender identity categories. Thirdly, we investigate to what extent possible gender differences in FUNCT and COFUNCT are explained by background and confounding factors related to FUNCT and COFUNCT. Finally, we investigate how background and confounding factors explain the variance of FUNCT and COFUNCT within the non-binary and binary participants.

## 2. Materials and methods

### 2.1. Participants and procedure

In May 2021, a Webropol survey was sent to the personnel and students of University of Turku, Finland. The survey was mailed to 27784 subjects (21227 students and 6557 personnel members) and re-mailed a week later. Study subjects responded anonymously. In all, 2004 subjects returned the survey and 1998 subjects (7.4%) completed the whole inquiry. The proportion of missing data was approximately 1% or less.

The survey assessed the following data: gender, background, living situation, adolescence school performance, friends, adversities, mental care, current contacts with family and friends, resilience, loneliness, depression symptoms, current functioning (FUNCT), and subjective evaluation of the effect of COVID-19 on functioning (COFUNCT).

### 2.2. Questions

The question assessing respondents’ gender included four options: 1) Female (Binary Female), 2) Male (Binary Male), 3) Other, 4) I do not wish to tell. In analyses, two latter options were combined to indicate non-binary gender. Other background factors included participant age, marital status, and living and work situation (employers, students and other [unemployed or sick]).

Functioning was assessed with two questions: “How is your ability to function?” and “How is your ability to work?” The visual analogue scale (VAS) (0 = very bad … 10 = very good) was used to collect responses. The VAS scores for these two questions were averaged to calculate FUNCT. Correspondingly, VAS scores for two additional questions, e.g., “How has the coronavirus epidemic affected your ability to function/work?” (Rated 0 = extremely negatively … 10 = extremely positively), were averaged to calculate COFUNCT.

School success and number of close friends at ages 12 to 18 were assessed with the VAS (0 = very badly/no friends at all … 10 = very well/a lot of close friends). Adverse childhood experiences (ACEs) were assessed with the Trauma and Distress Scale [25], which includes five core domains: emotional abuse, physical abuse, sexual abuse, emotional neglect and physical neglect.

Economic situation was assessed with a VAS question: “What is your current financial situation like?” and rated: 0 = extremely negative … 10 = extremely positive. Physical exercise was measured with the question: “How often do you exercise for at least half an hour at the time, so that you at least mildly get out of breath and sweat” (1 = daily, 2 = weekly, 3 = less often). Alcohol abuse was assessed using three questions: 1) “I think that I use too much alcohol”, 2) “My friend or family member has said that I use too much alcohol”, and 3) “I have sought professional help for my alcohol use and the problems it causes” (1 = Yes, 2 = No). Scores on these three questions were summed to calculate a total score for alcohol problems.

Family and friendship support comprised three questions: 1. “How many family members/friends do you see or hear from at least once a month?”, 2. “How many family members/friends do you feel at ease with that you can talk about private matters?”, 3. “How many family members/friends do you feel so close that you could call on them for help?” Each question was rated as follows: 1 = none, 2 = one, 3 = two, 4 = three to four and 5 = five or more. Sum of family and friends’ questions (range 3-15) indicated the level of support from family and friends.

Loneliness was calculated by summing the scores of three questions: “How often do you feel that you 1) lack companionship, 2) are left out, and 3) are isolated from others?” Each question was rated: 1 = hardly ever, 2 = some of the time, and 3 = often [26]. Resilience was assessed with two questions: 1) How quickly do you recover after adversity? and 2) If you fail, how long does your failure bother you? (0 = very slowly/for a very long time … 10 = very quickly/for a very short period). The VAS scores of these two questions were averaged to indicate resilience. Mental health care was assessed with a question “Have you, during the last 6 months, received care for mental health issues? (1 = Yes, 2 = No). Depressive symptoms (DEPs) were assessed with the DEPS scale [27].

### 2.3. Statistical analyses

Because we combined the participants who selected the gender options “Other” and “I do not wish to tell”, we first compared their characteristics. These analyses showed that participants who selected the latter option were older (mean years 34.9 vs. 26.7; p<0.001), had received mental health care less often (10.0% vs. 34.1%; p=0.018), and had better FUNCT scores (score 5.0 vs. 6.1; P=0.041) compared to the “Other” group. When the effect of age was controlled, there was no significant difference between these two non-binary groups (Supplementary Table 1). In univariate ANOVAs, both non-binary groups showed significant negative differences in FUNCT and COFUNCT (Supplementary Table 2). These analyses justify our decision to combine these two groups of participants as one non-binary group.

Distributions of background characteristics were cross-tabulated by gender and tested with the Chi-squared test (Table 1). Means (SD) for school success, friendships, ACEs, economy, support of family and friends, loneliness, resilience, depression, FUNCT, and COFUNCT were calculated and tested with ANOVA (Table 2). Pearson correlation coefficients were calculated (Supplementary Table 3). ANOVA was used to determine how FUNCT and COFUNCT were affected by background characteristics and continuous factors. In Model A, all background and confounding factors, except for depression, were used as independent explanators for FUNCT and COFUNCT. Only significant associations were reported (Table 3). In Model B, depression was included into independent factors, and only significant associations were reported (Table 4). In sensitivity ANOVA analyses for non-binary and binary participants separately, variance of FUNCT and COFUNCT was explained by all background and confounding factors (Supplementary Table 4 and 5). In ANOVAs, also estimated effect was reported (η2).

**Table 1.** Distribution of background characteristics by gender (%).

| Table 1. Distribution of background characteristics by gender (%). |  |  |  |  |  |  |  |
| --- | --- | --- | --- | --- | --- | --- | --- |
|  | NBG | BM | BF | All | NBG/BM | NBG/BF | BM/BF |
|  | n=71 | n=475 | n=1452 | n=1998 | p | p | p |
| Age |  |  |  |  | **<0.001** | **0.002** | **<0.001** |
| -20 | 11.3 | 2.1 | 5.3 | 4.8 |  |  |  |
| 21-30 | 56.3 | 45.5 | 50.4 | 49.4 |  |  |  |
| 31-40 | 22.5 | 20.4 | 17.9 | 18.7 |  |  |  |
| 41-50 | 5.6 | 14.9 | 13.6 | 13.7 |  |  |  |
| 51-60 | 4.2 | 9.7 | 10.1 | 9.8 |  |  |  |
| 61+ | 0.0 | 7.4 | 2.6 | 3.7 |  |  |  |
| Mean | 29.9 | 36.5 | 33.6 | 34.2 | **<0.001** | **0.014** | **<0.001** |
| Marital status |  |  |  |  | **0.004** | **0.002** | **0.004** |
| Single | 62.0 | 45.3 | 42.4 | 43.8 |  |  |  |
| Married | 9.9 | 31.6 | 25.7 | 26.5 |  |  |  |
| Cohabitation | 23.9 | 19.8 | 27.9 | 25.8 |  |  |  |
| Divorced | 2.8 | 2.9 | 3.8 | 3.6 |  |  |  |
| Widowed | 1.4 | 0.4 | 0.2 | 0.3 |  |  |  |
| Living with |  |  |  |  | **0.034** | **0.002** | 0.106 |
| Alone | 50.7 | 41.9 | 37.3 | 38.9 |  |  |  |
| Parents | 2.8 | 3.4 | 2.0 | 2.4 |  |  |  |
| Spouse/children | 35.2 | 49.5 | 54.3 | 52.5 |  |  |  |
| Other | 11.3 | 5.3 | 6.4 | 6.3 |  |  |  |
| Childhood family |  |  |  |  | **<0.001** | **0.005** | **0.007** |
| Two parents | 59.2 | 79.0 | 70.4 | 72.0 |  |  |  |
| One parent died | 0.0 | 3.0 | 3.9 | 3.5 |  |  |  |
| Parents divorced | 31.0 | 14.9 | 22.2 | 20.8 |  |  |  |
| One parent | 2.8 | 1.5 | 1.5 | 1.6 |  |  |  |
| Other | 7.0 | 1.5 | 2.0 | 2.1 |  |  |  |
| Work status |  |  |  |  | **0.004** | **0.001** | **0.002** |
| Employed | 25.4 | 44.8 | 38.1 | 39.2 |  |  |  |
| Student | 63.4 | 49.7 | 58.5 | 56.6 |  |  |  |
| Other | 11.3 | 5.5 | 3.4 | 4.2 |  |  |  |
| Mental health care; 6 months |  |  |  |  | 0.088 | 0.889 | **<0.001** |
| Yes | 23.9 | 15.6 | 25.1 | 22.8 |  |  |  |
| No | 76.1 | 84.4 | 74.9 | 77.2 |  |  |  |
| Alcohol abuse |  |  |  |  | 0.110 | 0.827 | **<0.001** |
| Yes | 8.5 | 16.0 | 8.2 | 10.1 |  |  |  |
| No | 91.5 | 84.0 | 91.8 | 89.9 |  |  |  |
| Physical exercise |  |  |  |  | **0.004** | **0.005** | 0.700 |
| Daily | 7.0 | 5.9 | 6.7 | 6.6 |  |  |  |
| Weekly | 56.3 | 74.3 | 72.5 | 72.4 |  |  |  |
| Less often | 36.6 | 19.8 | 20.7 | 21.1 |  |  |  |

NBG=Non-Binary Gender, BM=Binary Male, BF=Binary Female

**Table 2.** Means (SD) and ANOVAs of various characteristics by gender.

NBG=Non-Binary Gender, BM=Binary Male, BF=Binary Female
| Table 2. Means (SD) and ANOVAs of various characteristics by gender. |  |  |  |  |  |  |  |  |  |  |  |  |
| --- | --- | --- | --- | --- | --- | --- | --- | --- | --- | --- | --- | --- |
|  | NBG (n=71) |  | BM (n=475) |  | BF (n=1452) |  | All (n=1998) |  | ANOVA | ANOVA | ANOVA | ANOVA |
|  | Mean | SD | Mean | SD | Mean | SD | Mean | SD | NBG/BF/BM | NBG/BM | NBG/BF | BF/BM |
| Scholastic performance (0-10) | 8.085 | 1.722 | 7.935 | 2.066 | 8.532 | 1.627 | 8.374 | 1.763 | **<0.001** | 0.500 | **0.035** | **<0.001** |
| Friendships (0-10) | 5.141 | 2.748 | 5.634 | 2.821 | 6.046 | 2.762 | 5.915 | 2.784 | **0.001** | 0.163 | **0.007** | **0.005** |
| ACE sum | 20.676 | 16.823 | 11.051 | 9.479 | 12.130 | 10.672 | 12.177 | 10.811 | **<0.001** | **<0.001** | **<0.001** | 0.056 |
| Economy (0-10) | 5.324 | 2.329 | 6.442 | 2.523 | 6.304 | 2.441 | 6.302 | 2.464 | **0.002** | **<0.001** | **0.001** | 0.289 |
| Support from family (3-15) | 9.056 | 2.634 | 10.187 | 2.729 | 10.610 | 2.648 | 10.455 | 3.339 | **<0.001** | **0.001** | **<0.001** | **0.003** |
| Support from friends (3-15) | 10.000 | 3.692 | 10.145 | 3.536 | 11.023 | 3.222 | 10.778 | 3.339 | **<0.001** | 0.731 | **0.011** | **<0.001** |
| Loneliness (1-3) | 2.108 | 0.606 | 1.686 | 0.639 | 1.782 | 0.622 | 1.770 | 0.630 | **<0.001** | **<0.001** | **<0.001** | **0.004** |
| Resilience (0-10) | 4.338 | 1.822 | 5.792 | 2.119 | 5.138 | 2.011 | 5.265 | 2.056 | **<0.001** | **<0.001** | **0.001** | **<0.001** |
| Depression (2-40) | 24.296 | 7.322 | 18.634 | 6.604 | 19.592 | 6.818 | 19.532 | 6.856 | **<0.001** | **<0.001** | **<0.001** | **0.008** |
| Functioning (0-10) | 5.500 | 2.246 | 7.205 | 2.128 | 7.065 | 2.117 | 7.043 | 2.145 | **<0.001** | **<0.001** | **<0.001** | 0.212 |
| Effect of COVID-19 on functioning (0-10) | 3.500 | 1.371 | 4.737 | 1.752 | 4.713 | 1.846 | 4.675 | 1.823 | **<0.001** | **<0.001** | **<0.001** | 0.802 |

**Table 3.** ANOVA for functioning without depression and with depression. Statistically significant associations only. ACEs = Adverse childhood experiences, η2 = Eta squared.

|  | A. Without depression |  |  |  |  | B. With depression |  |  |  |  |
| --- | --- | --- | --- | --- | --- | --- | --- | --- | --- | --- |
| | B | p | CI95% | | $\eta^2$ | B | p | CI95% | | $\eta^2$ |
| Intercept | 5.950 | <0.001 | 5.419 | 6.482 | 0.195 | 8.913 | <0.001 | 8.444 | 9.383 | 0.411 |
| Gender |  | <0.001 |  |  |  |  | 0.010 |  |  |  |
| NBG | -0.744 | <0.001 | -1.145 | -0.343 | 0.007 | -0.473 | 0.007 | -0.817 | -0.128 | 0.004 |
| BM | -0.193 | 0.030 | -0.368 | -0.018 | 0.002 | -0.127 | 0.099 | -0.277 | 0.024 | 0.001 |
| BF | - |  |  |  |  | - |  |  |  |  |
| Childhood friends | -0.036 | 0.011 | -0.064 | -0.008 | 0.003 |  |  |  |  |  |
| ACEs | -0.013 | <0.001 | -0.021 | -0.006 | 0.006 |  |  |  |  |  |
| Work situation |  | <0.001 |  |  |  |  | <0.001 |  |  |  |
| Other | -1.254 | <0.001 | -1.650 | -0.859 | 0.019 | -0.973 | <0.001 | -1.315 | -0.631 | 0.015 |
| Student | -0.022 | 0.797 | -0.188 | 0.144 | 0.000 | 0.072 | 0.322 | -0.071 | 0.214 | 0.000 |
| Employed | - |  |  |  |  | - |  |  |  |  |
| Economy | 0.198 | <0.001 | 0.164 | 0.231 | 0.064 | 0.137 | <0.001 | 0.108 | 0.166 | 0.042 |
| Physical exercise | 0.189 | <0.001 | 0.133 | 0.244 | 0.022 | 0.103 | <0.001 | 0.055 | 0.152 | 0.009 |
| Loneliness | -0.697 | <0.001 | -0.830 | -0.563 | 0.050 |  |  |  |  |  |
| Resilience | 0.212 | <0.001 | 0.171 | 0.253 | 0.050 | 0.075 | <0.001 | 0.040 | 0.110 | 0.009 |
| Mental health care | -0.969 | <0.001 | -1.156 | -0.782 | 0.049 | -0.634 | <0.001 | -0.796 | -0.472 | 0.029 |
| Depression |  |  |  |  |  | -0.170 | <0.001 | -0.181 | -0.159 | 0.301 |
| $R^2 = 0.408$ (Adjusted $R^2 = 0.405$) | | | | | | $R^2 = 0.559$ (Adjusted $R^2 = 0.557$) | | | | |

NBG=Non-Binary Gender, BM=Binary Male, BF=Binary Female

**Table 4.** ANOVA for the effect of COVID-19 on functioning without depression and with depression. Statistically significant associations only. ACEs = Adverse childhood experiences, η2 = Eta squared

|  | A. Without depression |  |  |  |  | B. With depression |  |  |  |  |
| --- | --- | --- | --- | --- | --- | --- | --- | --- | --- | --- |
| | B | p | CI95% | | $\eta^2$ | B | p | CI95% | | $\eta^2$ |
| Intercept | 3.940 | <0.001 | 3.271 | 4.608 | 0.063 | 6.054 | <0.001 | 5.517 | 6.592 | 0.197 |
| Gender |  | <0.001 |  |  |  |  | 0.010 |  |  |  |
| NBG | -0.843 | <0.001 | -1.242 | -0.444 | 0.009 | -0.677 | 0.001 | -1.064 | -0.290 | 0.006 |
| BM | -0.239 | 0.008 | -0.415 | -0.063 | 0.004 | -0.167 | 0.052 | -0.335 | 0.002 | 0.002 |
| BF | - |  |  |  |  | - |  |  |  |  |
| Childhood friends | -0.031 | 0.038 | -0.060 | -0.002 | 0.002 |  |  |  |  |  |
| Age | 0.211 | <0.001 | 0.147 | 0.274 | 0.021 | 0.215 | <0.001 | 0.155 | 0.275 | 0.024 |
| Economy | 0.065 | <0.001 | 0.033 | 0.096 | 0.008 | 0.036 | 0.022 | 0.005 | 0.067 | 0.003 |
| Physical exercise | 0.107 | <0.001 | 0.051 | 0.162 | 0.007 | 0.070 | 0.012 | 0.016 | 0.125 | 0.003 |
| Support from family | 0.033 | 0.028 | 0.004 | 0.063 | 0.002 |  |  |  |  |  |
| Support from friends | -0.048 | <0.001 | -0.075 | -0.022 | 0.006 | -0.039 | 0.001 | -0.061 | -0.016 | 0.006 |
| Loneliness | -0.412 | <0.001 | -0.555 | -0.270 | 0.016 |  |  |  |  |  |
| Resilience | 0.130 | <0.001 | 0.089 | 0.171 | 0.019 |  |  |  |  |  |
| Mental care | -0.315 | 0.001 | -0.499 | -0.132 | 0.006 |  |  |  |  |  |
| Depression |  |  |  |  |  | -0.097 | <0.001 | -0.108 | -0.085 | 0.116 |
| $R^2 = 0.180$ (Adjusted $R^2 = 0.176$) | | | | | | $R^2 = 0.224$ (Adjusted $R^2 = 0.221$) | | | | |

NBG=Non-Binary Gender, BM=Binary Male, BF=Binary Female

Path analyses were performed using the PROCESS macro in SPSS (model template 4) by A. F. Hayes [28]. In cross-sectional samples, this macro tests the direct and indirect effects of an independent variable (X) on a dependent variable (Y) while modelling a process in which X affects a mediator (M), which in turn affects Y. The models tested the effect of ACEs/loneliness/resilience (X) on FUNCT/COFUNCT (Y) with depression as mediator (M). Total, direct and indirect effects via a mediator are reported (Supplementary Table 6).

The data were analysed using Statistical Program for the Social Sciences (SPSS) v28.0, and p values <0.05 (two-tailed) were considered statistically significant.

## 3. Results

### 3.1. Descriptive results

Of all 1998 study participants 41 (2.1%) selected the gender option “Other” and 30 (1.5%) the option “I do not want to tell”. Together, they form the group of non-binary participants (71, 3.6%). 452 (23.8%) participants selected the gender option “Male” and 1452 (72.7%) the option “Female”. Non-binary participants were younger and more likely to be single, live alone, and be unemployed or sick, compared to binary participants. Moreover, they practised physical exercise less often than binary participants. During their childhood, compared with binary participants, non-binary participants had lived less often with two parents and their parents had divorced more often (Table 1).

Compared with binary participants, non-binary participants reported more ACEs, a weaker economic status, less family support and resilience, and more loneliness and depression, as well as lower FUNCT and COFUNCT. In childhood, non-binary participants’ scholastic performance was poorer, and they had less friends than binary females (Table 2). In summary, in relation to binary participants, non-binary participants exhibited multiple social disadvantages.

FUNCT correlated strongly with ACEs, age, work situation, economy, physical exercise, family support, loneliness, resilience, mental health care and depression, while COFUNCT correlated with age, economy, loneliness, resilience and depression. Correlations related to FUNCT were stronger than those with COFUNCT (Supplementary Table 3).

### 3.2. Analyses of variance

#### 3.2.1. Functioning

In univariate ANOVA, FUNCT for non-binary participants (mean 5.500, SD 0.252, CI95% 5.006 to 6.994) was lower (p<0.001) than FUNCT in binary males (mean 7.205, SD 0.097, CI95% 7.014 to 7.396) and binary females (mean 7.065, SD 0.056. CI95% 6.956 to 7.174), but there was not statistically significant (p=0.212) difference between binary males and females.

In multivariate ANOVA for FUNCT, non-binary and binary male participants, numerous childhood friends (!), ACEs, poor work situation (unemployed/sick), loneliness, and mental health care associated with poor FUNCT, while good economy, physical exercise, and resilience associated with good FUNCT (Table 3, Model A). When depression was included into model, non-binary gender, poor work situation, mental health care, and depression associated with poor FUNCT, while good economy, physical exercise, and resilience associated with good FUNCT (Table 3, Model B). Of the ACE domains, emotional neglect (B: -0.032, p=0.002, CI95%: -0.012 to – 0.005) and physical neglect (B: -0.057, p<0.001, CI95%: -0.086 to – 0.027) separately associated with FUNCT. Emotional neglect associated with FUNCT only indirectly via depression, whereas physical neglect associated with FUNCT both directly and indirectly via depression.

In sensitivity analyses for non-binary participants, ACEs, poor work situation (unemployment or sick leave), and loneliness associated negatively with FUNCT. In binary participants, poor work situation, loneliness, and mental health care associated with poor FUNCT, but good economy, active physical exercise and good resilience associated with FUNCT. It was notable that among the non-binary participants, functioning supporting factors played no role (Supplementary Table 4). The effect of ACEs and loneliness on FUNCT was mediated via depression both in non-binary and binary participants (Supplementary Table 6).

#### 3.2.2. Effect of COVID-19 on functioning

In univariate ANOVA, non-binary participants reported more negative (p<0.001) COFUNCT (mean 3.500, SD 0.215, CI95% 3.079 to 3.921) than binary male (mean 4.737, SD 0.083, CI95% 4.574 to 4.900) and female participants (mean 4.713, SD 0.047. CI95% 4.620 to 4.806), but there was not statistically significant (p=0.802) difference between binary male and female participants.

In multivariate ANOVA, non-binary and binary male participants compared to binary females, ACEs, friends’ support (!), mental health care and loneliness associated with poor COFUNCT, while age, family support, economy, physical exercise, and resilience associated with good COFUNCT (Table 4, Model A). When depression was included in the model, non-binary gender, friends’ support (!), and depression associated with poor COFUNCT, whereas age, economy, physical exercise, and resilience associated with good COFUNCT (Table 4, Model B).

In sensitivity analyses for non-binary participants, family support (!), and loneliness associated negatively with COFUNCT. In binary participants, ACEs (!), age, economy, physical exercise, family support, and resilience associated positively, and friends’ support (!) and loneliness negatively with COFUNCT (Supplementary Table 5). In non-binary participants, childhood family associated statistically non-significantly (p=0.065) with COFUNCT. However, compared to those with two-parent families, participants whose parents were divorced reported more (mean difference 0.929, SD 0.390, p=0.021, CI95% 0.146 to 1.713) negative COFUNCT. When depression was included in the model, this difference between two-parent families and divorced-parent families was even greater (mean difference 1.006, SD 0.356, p=0.007, CI95% 0.290 to 1.721).

### 3.3. Path analyses

Univariate path analyses showed that, in the whole sample and in binary participants separately, ACEs and resilience had both direct and indirect effects, via depression, on FUNCT and COFUNCT. Loneliness had only an indirect effect on FUNCT and COFUNCT. In non-binary participants, ACEs, loneliness, and resilience had only an indirect effect on FUNCT and COFUNCT (Supplementary Table 6).

## 4. Discussion

### 4.1. Main results

Our main finding was that non-binary participants reported lower functioning and a greater negative effect of COVID-19 on their functioning compared to binary participants together or binary males and binary females separately. Hence, our main hypothesis was confirmed. Secondly, these gender differences mainly remained although the effects of background and confounding factors were considered. However, in multivariate analyses for FUNCT and COFUNCT, the binary males fared worse than binary females, possibly because gender-related factors that support FUNCT and COFUNCT, i.e., economy, physical exercise, and resilience, were statistically taken into account. Moreover, compared with binary participants, non-binary participants’ socioeconomic living situation and their previous psychosocial development was in many respects poorer indicating that compared with binary participants, non-binary participants had less positive functioning supporting psychosocial resources available during the societal stress related to COVID-19 pandemic.

### 4.2. Current functioning

In the present study, FUNCT represents the sustained ability to function, developed during individuals’ life-span, but it also includes effect of recent changes in functioning due to contemporary life-stress. ACEs played a major role in gender differences of functioning. Non-binary participants reported more ACEs, and their mental health was poorer than that of the binary participants, which explained a major part of FUNCT’s variance. Thus, compared with binary participants, non-binary participants’ lower FUNCT was developed over an extended period of time, possibly due to frequent ACEs that are generally associated with adult depression [25,29]. In univariate ANOVA, ACEs had both direct and indirect effects on FUNCT mediated through depression, while in multivariate analyses, the effect of ACEs on FUNCT was mediated via depression that was a major factor reducing FUNCT. Interestingly, in the binary participants ACEs had no significant, although indicative (p=0.057) association with FUNCT, possibly because of the effect of FUNCT supporting factors like economy, physical exercise, and resilience.

Of the ACEs, physical and emotional neglect were the main domains associating with decreased FUNCT, indicating that neglecting or disregarding children’s physical and emotional needs may lead to low motivation to act and achieve life goals, which manifests as lower functioning. The fact that physical neglect had both direct and indirect effect on FUNCT, may speak about model learned carelessness originating from the childhood family. In line with this social intergenerational transmission hypothesis [30], non-binary participants reported poorer economic situation, less resilience, and less physical exercise than binary participants indicating that their psychosocial resources supporting active functioning were consistently limited.

In line with previous studies, loneliness was more prevalent in non-binary participants [31,32], and associated strongly with FUNCT, and likewise in the case of ACEs, its effect on FUNCT was mediated via depression. It is noteworthy that although loneliness was strongly correlated with ACEs, it also had an independent association with FUNCT. In addition, adult loneliness not only correlated negatively with friends’ support, but it also showed a rather strong negative correlation (r=0.269) with childhood friends. This suggests that the roots of adult loneliness in non-binary participants extend back to early childhood, indicating that loneliness represents a persistent personality trait [33].

In the whole sample and in the binary participants separately, good economy, resilience, and active physical exercise associated with good FUNCT. However, in the non-binary participants, these factors played no role in explaining the variance of FUNCT. Thus, low levels of economy, resilience, and physical exercise in the non-binary participants could not support or maintain their ability to function. Finally, although the effects of background and confounding factors were controlled, non-binary participants still had poorer FUNCT than binary participants suggesting that there exist other factors associating with FUNCT, not considered in our models. For example, experiences of discrimination [5] may have diminished the psychological energy and daring for everyday functioning in non-binary individuals.

### 4.3. Effect of COVID-19 on functioning

COFUNCT indicates perceived changes in the functioning related to the current societal stress caused by the COVID-19 pandemic. This distinction explains some differences in ANOVA models. The fact that gender differences were mainly similar in both study approaches may be explained by the stronger gender effect on COFUNCT, that was partly included into FUNCT. In COFUNCT, ACEs and loneliness played a smaller role than in FUNCT, and as previously noted, their effect on COFUNCT was mediated via depression. This may indicate that prominent factors in non-binary individuals, such as ACEs and loneliness, make them vulnerable to depression. Additionally, due to a lack or insufficiency of stress-situation supporting factors, such as resilience and physical exercise, their ability to maintain everyday functioning suffers greatly in an exceptional stress situation.

Age associated strongly with COFUNCT: older participants reported a lesser negative effect of COVID-19 on their functioning. However, this association concerned only binary participants; age had no association with COFUNCT in non-binary participants. Thus, unlike the binary participants, increased life experience did not increase the non-binary participants’ resilience against societal stress. Probably, the early psychic vulnerability of non-binary individuals prevented their ability to build up strength for coping with various life stressors, leaving them unprepared and untrained to face exceptional societal stress situations.

Unexpectedly, among the binary participants, ACEs and family support associated positively with COFUNCT, while friends’ support showed negative association. It is possible that (moderate) childhood adversities, with increasing life experience and the protection of family support, binary individuals built up their strength and resources to manage exceptional life challenges like the COVID-19 pandemic and its social consequences, such like being forced to be separated from friends. In this process, when adverse childhood experiences turn to strengthening endurance or resilience, a good long-lasting family relationship may be decisive. In the non-binary participants, family situation was different: parents’ divorce and current family support associated negatively with COFUNCT, and after taking into account the effect of depression, these negative associations even strengthened. This means that not only ACEs and loneliness, but also longstanding family problems may have gnawed non-binary participants’ capability to get along with an exceptional social and societal stress.

### 4.4. Limitations

The present survey is a typical cross-sectional study and does not allow causal conclusions. We did not have knowledge on the participants’ functioning before the COVID-19 pandemic, which would clearly made comparison with current functioning more reliable. Therefore, the conclusions are mainly basing on the COFUNCT analyses. The low response rate (7.4%) also limits epidemiological representativeness of the results. The survey was not very long but it included also sensitive questions, which may have reduced individuals’ willingness to response. In client satisfaction surveys with no incentives, response rate often remains on the level of 10% or lower [34]. During the COVID-19 pandemic, the university students and personnel received several other surveys, thus it is probable that they were tired to response surveys. Additionally, the fact that this survey was carried out in May, when the term was near to its end, may have affected low response rate.

On the other hand, the sample size became large enough for studying also small groups of participants and possible associations between various factors and functioning. The prevalence (3.6%) of non-binary individuals received in the present study is small but comparable with the estimates reported in previous studies [4,35,36]. Yet, the low number of non-binary participants puts some limitations for more detailed analyses.

The study focused on people of university community, who do not represent the general population. On the other hand, the study sample represents a quite homogenous population, which faced equal and long-lasting COVID-19 lockdown with its stressful consequences.

The question regarding gender provided only four options (Female, Male, Other, I do not wish to tell) and all except reported females and males were combined to one group of non-binary gender. Although, the statistical analyses indicated that the two non-binary groups were comparable, it is possible that the reasons why some participants selected the option “I do not wish to tell” was not related to their gender identity. Anyway, the non-binary group is heterogeneous group including several types of genderqueers and individuals who may be afraid that they can be traced from their responses. Therefore, results related to gender should be regarded provisionally, not conclusive; more detailed studies are needed.

## 5. Conclusions

In the university community, compared with the individuals with binary gender identity, the non-binary individuals are more vulnerable for functional deficits during societal stress such as a COVID-19-pandemic. Psychosocial factors such as parents’ divorce, other childhood adversities, adult loneliness, and mental health problems are associated with lower functioning in these individuals. These characteristics should be taken into account when planning interventions in vulnerable groups including individuals with non-binary gender.

## 6. Additional information

### 1. Ethics approval and consent to participate

The study is carried out according to the Helsinki Declaration. The ethical committee of University of Turku approved the study protocol. All study subjects gave their written consent to the study.

### 3. Availability of data and materials

According to the decision of the ethical committee, the data can be used only for this study and are not available to other parties.

