## Supplementary Material for "Non-binary gender identity associates with decreased functioning during an exceptional societal stress. A university community study during COVID-19 pandemic in Finland"

Supplementary table 1. Comparisons between non-binary participant groups: Option 3 "Other" and Option 4: "I do not wish to tell". ACEs=Adverse childhood experiences

| | Mean | SD | Mean | SD | Mean | SD | p1 | p2 |
|---|---|---|---|---|---|---|---|---|
| Age | 26.707 | 5.853 | 34.267 | 10.034 | 29.901 | 8.685 | <0.001 | |
| Scholastic performance | 8.317 | 1.823 | 7.767 | 1.547 | 8.085 | 1.722 | 0.185 | 0.340 |
| Friendships | 5.073 | 2.841 | 5.233 | 2.661 | 5.141 | 2.748 | 0.810 | 0.705 |
| ACE sum | 19.073 | 15.342 | 22.867 | 18.706 | 20.676 | 16.823 | 0.352 | 0.491 |
| Economy | 5.146 | 2.319 | 5.567 | 2.359 | 5.324 | 2.329 | 0.456 | 0.934 |
| Support from family | 9.146 | 2.163 | 8.933 | 3.205 | 9.056 | 2.634 | 0.739 | 0.899 |
| Support from friends | 10.732 | 3.471 | 9.000 | 3.806 | 10.000 | 3.692 | 0.050 | 0.103 |
| Physical exercise | 3.195 | 1.364 | 3.633 | 1.650 | 3.380 | 1.496 | 0.225 | 0.141 |
| Mental health care | 0.342 | 0.480 | 0.100 | 0.305 | 0.239 | 0.430 | 0.018 | 0.055 |
| Loneliness | 2.146 | 0.573 | 2.056 | 0.656 | 2.108 | 0.606 | 0.537 | 0.637 |
| Resilience | 4.122 | 1.867 | 4.633 | 1.747 | 4.338 | 1.822 | 0.246 | 0.691 |
| Depression | 24.951 | 7.152 | 23.400 | 7.578 | 24.296 | 7.322 | 0.382 | 0.869 |
| Functioning | 5.037 | 2.420 | 6.133 | 1.838 | 5.500 | 2.246 | 0.041 | 0.101 |
| Effect of COVID-19 on functioning | 3.342 | 1.362 | 3.717 | 1.375 | 3.500 | 1.371 | 0.257 | 0.455 |

p1 = significance between non-binary groups; p2 = significance between non-binary group when the effect of age is controlled.

| Supplementary table 2. ANOVA for functioning and for the effect of the COVID-19 pandemic on functioning in binary females, males and in non-binary participants (the participants who selected the gender option "Other" or "I do wish to tell") separately. | | | | | |
|---|---|---|---|---|---|
| | Functioning | | | | |
| | B | p | CI95% | | η2 |
| Intercept | 7.065 | <0.001 | 6.956 | 7.174 | 0.890 |
| Non-binary group (N=71) | -1.565 | <0.001 | -2.071 | -1.059 | 0.018 |
| Binary males (=475) | 0.140 | 0.212 | -0.080 | 0.360 | 0.001 |
| Binary females (N=1452) | - | | | | |
| | B | p | CI95% | | η2 |
| Intercept | 7.065 | <0.001 | 6.956 | 7.175 | 0.891 |
| "Other" (N=41) | -2.028 | <0.001 | -2.689 | -1.368 | 0.018 |
| Binary males (=475) | 0.140 | 0.212 | -0.080 | 0.361 | 0.001 |
| Binary females (N=1452) | - | | | | |
| | B | p | CI95% | | η2 |
| Intercept | 7.065 | <0.001 | 6.956 | 7.174 | 0.892 |
| "I do not wish to tell" (N=30) | -0.932 | 0.017 | -1.697 | -0.166 | 0.003 |
| Binary males (=475) | 0.140 | 0.210 | -0.079 | 0.360 | 0.001 |
| Binary females (N=1452) | - | | | | |
| | Effect of COVID-19 of functioning | | | | |
| | B | p | CI95% | | η2 |
| Intercept | 4.713 | <0.001 | 4.620 | 4.806 | 0.832 |
| Non-binary group (N=71) | -1.213 | <0.001 | -1.644 | -0.781 | 0.015 |
| Binary males (=475) | 0.024 | 0.802 | -0.164 | 0.212 | 0.000 |
| Binary females (N=1452) | - | | | | |
| | B | p | CI95% | | η2 |
| Intercept | 4.713 | <0.001 | 4.619 | 4.806 | 0.833 |
| "Other" (N=41) | -1.371 | <0.001 | -1.935 | -0.808 | 0.011 |
| Binary males (=475) | 0.024 | 0.802 | -0.164 | 0.212 | 0.000 |
| Binary females (N=1452) | - | | | | |
| | B | p | CI95% | | η2 |
| Intercept | 4.713 | <0.001 | 4.619 | 4.806 | 0.833 |
| "I do not wish to tell" (N=30) | -0.996 | 0.003 | -1.654 | -0.339 | 0.004 |
| Binary males (=475) | 0.024 | 0.803 | -0.164 | 0.212 | 0.000 |
| Binary females (N=1452) | - | | | | |

Supplementary table 3. Pearson's correlation coefficients for explanatory factors and functioning (FUNCT) and effect of COVID-19 on functioning (COFUNCT).

| Correlations | 1. | 2. | 3. | 4. | 5. | 6. | 7. | 8. | 9. | 10. | 11. | 12. | 13. | 14. | 15. | 16. | 17. | 18. | 19. | 20. |
|---|---|---|---|---|---|---|---|---|---|---|---|---|---|---|---|---|---|---|---|---|
| 1. Gender | 1 | -,048* | 0,032 | ,053* | -,151* | ,052* | ,070* | ,047* | -,068* | ,076* | ,045* | 0,01 | ,100* | ,045* | -,103* | ,087* | 0,005 | -,133* | ,138* | ,124* |
| 2. Childhood family | -,048* | 1 | -0,033 | -0,001 | ,226* | -,077* | -0,007 | -0,013 | 0,007 | -,107* | -0,019 | 0,031 | -,126* | 0,038 | 0,032 | -0,022 | -,072* | ,070* | -,081* | -,055* |
| 3. School success | 0,032 | -0,033 | 1 | ,176* | -,182* | -,098* | -0,024 | 0,024 | -0,018 | ,143* | ,048* | -,058* | ,088* | ,132* | -,063* | 0,012 | -0,03 | -,057* | ,090* | 0,002 |
| 4. Friends | ,053* | -0,001 | ,176* | 1 | -,186* | 0,005 | ,096* | ,074* | -0,03 | ,067* | ,097* | -0,007 | ,223* | ,374* | -,269* | ,243* | 0,039 | -,141* | ,113* | 0,029 |
| 5. ACEs | -,151* | ,226* | -,182* | -,186* | 1 | 0,009 | -0,007 | -0,021 | ,127* | -,201* | -,084* | ,120* | -,405* | -,188* | ,299* | -,267* | -,227* | ,341* | -,295* | -,099* |
| 6. Age | ,052* | -,077* | -,098* | 0,005 | 0,009 | 1 | ,422* | ,297* | 0,013 | ,245* | ,061* | ,055* | ,072* | -,203* | -,235* | ,233* | ,176* | -,261* | ,202* | ,276* |
| 7. Marital status | ,070* | -0,007 | -0,024 | ,096* | -0,007 | ,422* | 1 | ,780* | -0,001 | ,148* | 0,042 | 0,014 | ,220* | -,061* | -,173* | ,145* | ,128* | -,194* | ,160* | ,159* |
| 8. Living situation | ,047* | -0,013 | 0,024 | ,074* | -0,021 | ,297* | ,780* | 1 | 0,009 | ,125* | 0,028 | -0,016 | ,221* | -,063* | -,144* | ,138* | ,119* | -,172* | ,140* | ,152* |
| 9. Work situation | -,068* | 0,007 | -0,018 | -0,03 | ,127* | 0,013 | -0,001 | 0,009 | 1 | -,144* | -,049* | 0,021 | -,075* | -,062* | ,094* | -,082* | -,207* | ,159* | -,243* | -,054* |
| 10. Economy | ,076* | -,107* | ,143* | ,067* | -,201* | ,245* | ,148* | ,125* | -,144* | 1 | ,113* | -0,012 | ,162* | 0,042 | -,227* | ,230* | ,158* | -,341* | ,396* | ,217* |
| 11. Physical exercise | ,045* | -0,019 | ,048* | ,097* | -,084* | ,061* | 0,042 | 0,028 | -,049* | ,113* | 1 | 0 | ,125* | ,116* | -,177* | ,157* | ,078* | -,235* | ,236* | ,146* |
| 12. Alcohol abuse | 0,01 | 0,031 | -,058* | -0,007 | ,120* | ,055* | 0,014 | -0,016 | 0,021 | -0,012 | 0 | 1 | -0,043 | -0,002 | ,061* | -0,043 | -0,036 | ,094* | -,060* | -0,009 |
| 13. Support from family | ,100* | -,126* | ,088* | ,223* | -,405* | ,072* | ,220* | ,221* | -,075* | ,162* | ,125* | -0,043 | 1 | ,278* | -,309* | ,253* | ,166* | -,328* | ,251* | ,154* |
| 14. Support from friens | ,045* | 0,038 | ,132* | ,374* | -,188* | -,203* | -,061* | -,063* | -,062* | 0,042 | ,116* | -0,002 | ,278* | 1 | -,369* | ,148* | -0,01 | -,165* | ,111* | -0,027 |
| 15. Loneliness | -,103* | 0,032 | -,063* | -,269* | ,299* | -,235* | -,173* | -,144* | ,094* | -,227* | -,177* | ,061* | -,309* | -,369* | 1 | -,391* | -,209* | ,582* | -,419* | -,258* |
| 16. Resilience | ,087* | -0,022 | 0,012 | ,243* | -,267* | ,233* | ,145* | ,138* | -,082* | ,230* | ,157* | -0,043 | ,253* | ,148* | -,391* | 1 | ,246* | -,486* | ,418* | ,275* |
| 17. Mental health care | 0,005 | -,072* | -0,03 | 0,039 | -,227* | ,176* | ,128* | ,119* | -,207* | ,158* | ,078* | -0,036 | ,166* | -0,01 | -,209* | ,246* | 1 | -,324* | ,362* | ,187* |
| 18. Depression | -,133* | ,070* | -,057* | -,141* | ,341* | -,261* | -,194* | -,172* | ,159* | -,341* | -,235* | ,094* | -,328* | -,165* | ,582* | -,486* | -,324* | 1 | -,702* | -,426* |
| 19. FUNCT | ,138* | -,081* | ,090* | ,113* | -,295* | ,202* | ,160* | ,140* | -,243* | ,396* | ,236* | -,060* | ,251* | ,111* | -,419* | ,418* | ,362* | -,702* | 1 | ,512* |
| 20. COFUNCT | ,124* | -,055* | 0,002 | 0,029 | -,099* | ,276* | ,159* | ,152* | -,054* | ,217* | ,146* | -0,009 | ,154* | -0,027 | -,258* | ,275* | ,187* | -,426* | ,512* | 1 |

Gender: Non-binary gender/binary gender; Two parents/Other; ACEs: Childhood adversities; Marital status: Single/Ever married or cohabitant; Living situation: Alone/Other; Work situation: Employed or Student/Other; Mental health care during past 6 months: Yes/No; * Correlation is significant on p<0,05 level (2-tailed). ACEs=Adverse childhood experiences.

Supplementary table 4. ANOVAs for functioning in non-binary (NBG) and binary (BM+BF) gender, participants without depression and with depression. ACEs=Adverse childhood experiences. MSq=Mean squared. η2=Eta squared. Significant associations **bolded.**

| | NBG | | | | BM+BF | | | |
|---|---|---|---|---|---|---|---|---|
| **Without depression** | MSq | F | p | η2 | MSq | F | p | η2 |
| Corrected Model | **9.301** | **2.890** | **0.001** | **0.553** | **157.521** | **57.778** | **0.000** | **0.400** |
| Intercept | **30.854** | **9.588** | **0.003** | **0.164** | **279.192** | **102.406** | **0.000** | **0.051** |
| Gender | . | . | . | 0.000 | 7.874 | 2.888 | 0.089 | 0.002 |
| Childhood family | 2.826 | 0.878 | 0.422 | 0.035 | 2.441 | 0.895 | 0.409 | 0.001 |
| School success | 0.636 | 0.198 | 0.659 | 0.004 | 10.092 | 3.702 | 0.055 | 0.002 |
| Friends | 2.430 | 0.755 | 0.389 | 0.015 | 10.762 | 3.948 | 0.047 | 0.002 |
| ACEs | **16.264** | **5.054** | **0.029** | **0.093** | 9.865 | 3.618 | 0.057 | 0.002 |
| Age | 0.004 | 0.001 | 0.972 | 0.000 | 0.019 | 0.007 | 0.934 | 0.000 |
| Marital status | 1.622 | 0.504 | 0.607 | 0.020 | 0.952 | 0.349 | 0.705 | 0.000 |
| Living situation | 1.884 | 0.585 | 0.627 | 0.035 | 0.548 | 0.201 | 0.896 | 0.000 |
| Work situation | **15.696** | **4.877** | **0.012** | **0.166** | **48.659** | **17.848** | **0.000** | **0.018** |
| Economy | 3.206 | 0.996 | 0.323 | 0.020 | **344.501** | **126.361** | **0.000** | **0.062** |
| Physical exercise | 0.581 | 0.180 | 0.673 | 0.004 | **124.763** | **45.762** | **0.000** | **0.023** |
| Alcohol abuse | 8.255 | 2.565 | 0.116 | 0.050 | 1.999 | 0.733 | 0.392 | 0.000 |
| Support from family | 3.122 | 0.970 | 0.330 | 0.019 | 4.930 | 1.808 | 0.179 | 0.001 |
| Support from friends | 0.103 | 0.032 | 0.859 | 0.001 | 9.384 | 3.442 | 0.064 | 0.002 |
| Loneliness | **30.645** | **9.523** | **0.003** | **0.163** | **240.803** | **88.325** | **0.000** | **0.044** |
| Resilience | 1.562 | 0.485 | 0.489 | 0.010 | **275.654** | **101.109** | **0.000** | **0.050** |
| Mental health care | 8.664 | 2.692 | 0.107 | 0.052 | **254.941** | **93.511** | **0.000** | **0.047** |
| | $R^2$ = 0.553 (Adjusted $R^2$ = 0.362) | | | | $R^2$ = 0.400 (Adjusted $R^2$ = 0.393) | | | |
| | NBG | | | | BM+BF | | | |
| **With depression** | MSq | F | p | η2 | MSq | F | p | η2 |
| Corrected Model | **11.168** | **4.995** | **<0.001** | **0.696** | **207.870** | **102.075** | **<0.001** | **0.552** |
| Intercept | **55.236** | **24.707** | **<0.001** | **0.340** | **837.861** | **411.433** | **<0.001** | **0.178** |
| Gender | . | . | . | 0.000 | 5.201 | 2.554 | 0.110 | 0.001 |
| Childhood family | 4.030 | 1.802 | 0.176 | 0.070 | 1.683 | 0.827 | 0.438 | 0.001 |
| School success | 1.179 | 0.527 | 0.471 | 0.011 | **8.463** | **4.156** | **0.042** | **0.002** |
| Friends | 0.695 | 0.311 | 0.580 | 0.006 | 0.100 | 0.049 | 0.824 | 0.000 |
| ACEs | 2.333 | 1.043 | 0.312 | 0.021 | 0.006 | 0.003 | 0.958 | 0.000 |
| Age | 6.129 | 2.741 | 0.104 | 0.054 | 1.633 | 0.802 | 0.371 | 0.000 |
| Marital status | 2.276 | 1.018 | 0.369 | 0.041 | 1.140 | 0.560 | 0.571 | 0.001 |
| Living situation | 2.339 | 1.046 | 0.381 | 0.061 | 0.324 | 0.159 | 0.924 | 0.000 |
| Work situation | **10.575** | **4.730** | **0.013** | **0.165** | **31.906** | **15.667** | **<0.001** | **0.016** |
| Economy | 0.543 | 0.243 | 0.624 | 0.005 | **159.544** | **78.344** | **<0.001** | **0.040** |
| Physical exercise | 1.660 | 0.743 | 0.393 | 0.015 | **38.937** | **19.120** | **<0.001** | **0.010** |
| Alcohol abuse | 2.166 | 0.969 | 0.330 | 0.020 | 0.925 | 0.454 | 0.500 | 0.000 |
| Support from family | 4.093 | 1.831 | 0.182 | 0.037 | 0.891 | 0.438 | 0.508 | 0.000 |
| Support from friends | 0.754 | 0.337 | 0.564 | 0.007 | 2.409 | 1.183 | 0.277 | 0.001 |
| Loneliness | 3.557 | 1.591 | 0.213 | 0.032 | 0.060 | 0.030 | 0.863 | 0.000 |
| Resilience | 1.777 | 0.795 | 0.377 | 0.016 | **42.871** | **21.052** | **<0.001** | **0.011** |
| Mental health care | **12.193** | **5.454** | **0.024** | **0.102** | **104.278** | **51.206** | **<0.001** | **0.026** |
| Depression | **50.375** | **22.533** | **<0.001** | **0.319** | **1315.551** | **646.003** | **<0.001** | **0.253** |
| | $R^2$ = 0.696 (Adjusted $R^2$ = 0.557) | | | | $R^2$ = 0.552 (Adjusted $R^2$ = 0.547) | | | |

NBG=Non-Binary Gender, BM=Binary Male, BF=Binary Female

Table 5. ANOVAs for the effect of COVID-19 on functioning in non-binary (NBG) and binary (BG) gender participants. ACEs=Adverse childhood experiences. MSq=Mean squared. η2=Eta squared. Significant associations **bolded.**

| | NBG | | | | BG(BM+BF) | | | |
|---|---|---|---|---|---|---|---|---|
| **Without depression** | MSq | F | p | η2 | MSq | F | p | η2 |
| Intercept | **13.946** | **8.732** | **0.005** | **0.151** | **172.308** | **62.017** | **<0.001** | **0.032** |
| Gender | . | . | . | 0.000 | **18.179** | **6.543** | **0.011** | **0.003** |
| Childhood family | 4.622 | 2.894 | 0.065 | 0.106 | 2.203 | 0.793 | 0.453 | 0.001 |
| School success | 1.899 | 1.189 | 0.281 | 0.024 | 0.057 | 0.021 | 0.886 | 0.000 |
| Friends | 0.853 | 0.534 | 0.468 | 0.011 | 7.121 | 2.563 | 0.110 | 0.001 |
| ACEs | 3.572 | 2.236 | 0.141 | 0.044 | **12.087** | **4.350** | **0.037** | **0.002** |
| Age | 0.279 | 0.175 | 0.678 | 0.004 | **59.902** | **21.560** | **<0.001** | **0.011** |
| Marital status | 0.037 | 0.023 | 0.977 | 0.001 | 1.637 | 0.589 | 0.555 | 0.001 |
| Living situation | 0.871 | 0.545 | 0.654 | 0.032 | 3.052 | 1.098 | 0.349 | 0.002 |
| Work situation | 0.523 | 0.328 | 0.722 | 0.013 | 0.025 | 0.009 | 0.991 | 0.000 |
| Economy | 1.150 | 0.720 | 0.400 | 0.014 | **47.098** | **16.952** | **<0.001** | **0.009** |
| Physical exercise | 0.072 | 0.045 | 0.833 | 0.001 | **41.201** | **14.829** | **<0.001** | **0.008** |
| Alcohol abuse | 2.336 | 1.463 | 0.232 | 0.029 | 0.168 | 0.060 | 0.806 | 0.000 |
| Support from family | **6.783** | **4.247** | **0.045** | **0.080** | **20.135** | **7.247** | **0.007** | **0.004** |
| Support from friends | 0.138 | 0.087 | 0.770 | 0.002 | **36.638** | **13.187** | **<0.001** | **0.007** |
| Loneliness | **13.017** | **8.150** | **0.006** | **0.143** | **84.717** | **30.491** | **<0.001** | **0.016** |
| Resilience | 1.150 | 0.720 | 0.400 | 0.014 | **107.102** | **38.548** | **<0.001** | **0.020** |
| Mental health care | 4.287 | 2.684 | 0.108 | 0.052 | **29.081** | **10.467** | **0.001** | **0.005** |
| | $R^2$ = 0.405 (Adjusted $R^2$ = 0.150) | | | | $R^2$ = 0.174 (Adjusted $R^2$ = 0.164) | | | |
| | NBG | | | | BM+BF | | | |
| **With depression** | MSq | F | p | η2 | MSq | F | p | η2 |
| Intercept | **22.209** | **16.757** | **<0.001** | **0.259** | **359.330** | **137.967** | **<0.001** | **0.068** |
| Gender | . | . | . | 0.000 | **15.990** | **6.139** | **0.013** | **0.003** |
| Childhood family | **5.398** | **4.073** | **0.023** | **0.145** | 2.539 | 0.975 | 0.377 | 0.001 |
| School success | 2.350 | 1.773 | 0.189 | 0.036 | 0.140 | 0.054 | 0.817 | 0.000 |
| Friends | 0.283 | 0.214 | 0.646 | 0.004 | 1.374 | 0.528 | 0.468 | 0.000 |
| ACEs | 0.307 | 0.232 | 0.633 | 0.005 | **25.030** | **9.611** | **0.002** | **0.005** |
| Age | 0.649 | 0.490 | 0.487 | 0.010 | **49.293** | **18.926** | **<0.001** | **0.010** |
| Marital status | 0.087 | 0.066 | 0.936 | 0.003 | 1.681 | 0.646 | 0.525 | 0.001 |
| Living situation | 0.915 | 0.691 | 0.562 | 0.041 | 3.897 | 1.496 | 0.214 | 0.002 |
| Work situation | 0.088 | 0.066 | 0.936 | 0.003 | 0.313 | 0.120 | 0.887 | 0.000 |
| Economy | 2.620 | 1.976 | 0.166 | 0.040 | **15.264** | **5.861** | **0.016** | **0.003** |
| Physical exercise | 0.305 | 0.230 | 0.634 | 0.005 | **15.434** | **5.926** | **0.015** | **0.003** |
| Alcohol abuse | 0.598 | 0.451 | 0.505 | 0.009 | 2.571 | 0.987 | 0.321 | 0.001 |
| Support from family | **7.516** | **5.671** | **0.021** | **0.106** | 8.296 | 3.185 | 0.074 | 0.002 |
| Support from friends | 0.444 | 0.335 | 0.565 | 0.007 | **27.961** | **10.736** | **0.001** | **0.006** |
| Loneliness | 2.489 | 1.878 | 0.177 | 0.038 | 1.319 | 0.507 | 0.477 | 0.000 |
| Resilience | 0.120 | 0.090 | 0.765 | 0.002 | **27.197** | **10.442** | **0.001** | **0.005** |
| Mental health care | **5.592** | **4.219** | **0.045** | **0.081** | 6.376 | 2.448 | 0.118 | 0.001 |
| Depression | **14.648** | **11.052** | **0.002** | **0.187** | **333.789** | **128.160** | **<0.001** | **0.063** |
| | $R^2$ = 0.516 (Adjusted $R^2$ = 0.294) | | | | $R^2$ = 0.226 (Adjusted $R^2$ = 0.216) | | | |

NBG=Non-Binary Gender, BM=Binary Male, BF=Binary Female

Table 6. Path analyses for the direct and indirect, via depression, effects of Adverse Childhood Experiences (ACEs), loneliness, and resilience on functioning (FUNCT) and on the effect of COVID-19 on functioning (COFUNCT). Significant association **bolded. NBG =** non-binary gender**, BG =** binary gender.

| **All** | FUNCT | | | | COFUNCT | | | |
|---|---|---|---|---|---|---|---|---|
| ACEs | Effect | p | CI95% | | Effect | p | CI95% | |
| Total | **-0.059** | **<0.001** | **-0.067** | **-0.050** | **-0.017** | **<0.001** | **-0.024** | **-0.009** |
| Direct | **-0.012** | **<0.001** | **-0.019** | **-0.006** | **0.009** | **0.014** | **0.002** | **0.016** |
| Indirect | **-0.046** | | **-0.053** | **-0.039** | **-0.026** | | **-0.030** | **-0.022** |
| Loneliness | Effect | p | CI95% | | Effect | p | CI95% | |
| Total | **-1.428** | **<0.001** | **-1.563** | **-1.292** | **-0.747** | **<0.001** | **-0.870** | **-0.625** |
| Direct | -0.055 | 0.413 | -0.185 | 0.076 | -0.045 | 0.528 | -0.187 | 0.096 |
| Indirect | **-1.373** | | **-1.493** | **-1.259** | **-0.702** | | **-0.794** | **-0.612** |
| Resilience | Effect | p | CI95% | | Effect | p | CI95% | |
| Total | **0.436** | **<0.001** | **0.394** | **0.478** | **0.243** | **<0.001** | **0.206** | **0.281** |
| Direct | **0.105** | **<0.001** | **0.068** | **0.142** | **0.078** | **<0.001** | **0.038** | **0.119** |
| Indirect | **0.331** | | **0.298** | **0.366** | **0.165** | | **0.142** | **0.189** |
| **NBG** | FUNCT | | | | COFUNCT | | | |
| ACEs | Effect | p | CI95% | | Effect | p | CI95% | |
| Total | **-0.045** | **0.004** | **-0.075** | **-0.014** | -0.018 | 0.059 | -0.037 | 0.001 |
| Direct | -0.015 | 0.254 | -0.040 | 0.011 | -0.004 | 0.673 | -0.022 | 0.014 |
| Indirect | **-0.030** | | **-0.057** | **-0.006** | **-0.015** | | **-0.031** | **-0.002** |
| Loneliness | Effect | p | CI95% | | Effect | p | CI95% | |
| Total | **-1.419** | **0.001** | **-2.240** | **-0.597** | **-0.719** | **0.007** | **-1.234** | **-0.204** |
| Direct | -0.199 | 0.613 | -0.983 | 0.584 | -0.160 | 0.562 | -0.707 | 0.388 |
| Indirect | **-1.219** | | **-1.825** | **-0.690** | **-0.559** | | **-0.990** | **-0.225** |
| Resilience | Effect | p | CI95% | | Effect | p | CI95% | |
| Total | **0.324** | **0.027** | **0.038** | **0.609** | **0.175** | **0.050** | **0.000** | **0.351** |
| Direct | -0.012 | 0.925 | -0.256 | 0.233 | 0.021 | 0.806 | -0.150 | 0.192 |
| Indirect | **0.335** | | **0.147** | **0.542** | **0.154** | | **0.061** | **0.260** |
| **BG** | FUNCT | | | | COFUNCT | | | |
| ACEs | Effect | p | CI95% | | Effect | p | CI95% | |
| Total | **-0.057** | **<0.001** | **-0.065** | **-0.048** | **-0.013** | **0.001** | **-0.021** | **0.006** |
| Direct | **-0.011** | **0.001** | **-0.018** | **0.004** | **0.012** | **0.002** | **0.004** | **0.019** |
| Indirect | **-0.045** | | **-0.052** | **-0.039** | **-0.025** | | **-0.030** | **-0.021** |
| Loneliness | Effect | p | CI95% | | Effect | p | CI95% | |
| Total | **-1.393** | **<0.001** | **-1.531** | **-1.256** | **-0.718** | **<0.001** | **-0.844** | **-0.592** |
| Direct | -0.042 | 0.534 | -0.175 | 0.091 | -0.032 | 0.662 | -0.177 | 0.113 |
| Indirect | **-1.351** | | **-1.472** | **-1.223** | **-0.686** | | **-0.783** | **-0.592** |
| Resilience | Effect | p | CI95% | | Effect | p | CI95% | |
| Total | **0.430** | **<0.001** | **0.388** | **0.471** | **0.237** | **<0.001** | **0.199** | **0.276** |
| Direct | **0.107** | **<0.001** | **0.069** | **0.144** | **0.078** | **<0.001** | **0.037** | **0.119** |
| Indirect | **0.323** | | **0.290** | **0.356** | **0.159** | | **0.136** | **0.183** |

NBG=Non-Binary Gender, BM=Binary Male, BF=Binary Female
